# Influenza virus circulation and vaccine effectiveness during June 2021–May 2023 in Thailand

**DOI:** 10.1101/2023.09.29.23296370

**Authors:** Kriengkrai Prasert, Prabda Praphasiri, Sutthichai Nakphook, Darunee Ditsungnoen, Patranuch Sapchookul, Kanlaya Sornwong, Suriya Naosri, Pilailuk Akkapaiboon Okada, Piyarat Suntarattiwong, Tawee Chotpitayasunondh, Martha P. Montgomery, William W. Davis, Chakrarat Pittayawonganon

## Abstract

We evaluated 2023 Southern Hemisphere influenza vaccine effectiveness against medically attended illness using surveillance data from nine Thai hospitals and a test-negative design. During June 2022–May 2023, influenza vaccine provided protection against seeking care for influenza illness (adjusted vaccine effectiveness 43%; 95% confidence interval 15–61).

---

Influenza contributes substantially to morbidity and mortality in Thailand and worldwide.(1) Thai Ministry of Public Health recommends influenza vaccination for pregnant people, health care workers, adults aged ≥65 years, children aged 6–35 months, and people with underlying medical conditions. Vaccination using a Southern Hemisphere formulation is typically offered starting in May. Monitoring influenza activity can inform vaccination timing. Estimating vaccine effectiveness (VE) can help guide future antigen selection and reassure clinicians to continue to make a strong influenza vaccine recommendation to patients. This report describes temporal trends of influenza virus infection and influenza VE during two seasons in Thailand.

Influenza sentinel surveillance was established in Thailand in 2005.(2, 3) Nine hospitals in eight provinces representing major regions of Thailand conducted influenza-like illness (ILI) surveillance in their clinics and severe acute respiratory infection (SARI) surveillance in their wards. We defined ILI as measured fever ≥ 38 C and cough with onset within 10 days, and we defined SARI as fever history or measured fever ≥ 38 C and cough with onset within 10 days requiring hospitalization. Sites enrolled a convenience sample of two ILI patients (one age <5 and one ≥ 5 years) and two SARI patients (one age <5 and one ≥ 5 years) per working day for up to 20 patients total per week. Demographic, clinical, and self-reported influenza vaccination data were collected on a standardized form. Data were collected as routine public health surveillance, and written consent was not obtained. This project was reviewed by the Centers for Disease Control and Prevention (CDC) and was conducted consistent with applicable federal law and CDC policy (e.g., 45 CFR 46.102(l)(2)).

Nasopharyngeal or throat swab specimens were stored in viral transport media and tested at each site by QIAstat-Dx analyzer. This real-time reverse transcriptase polymerase chain reaction detected 19 viral and 3 bacterial pathogen targets, including influenza A, influenza A/H1N12009, influenza A H1, influenza A H3, and influenza B virus. We conducted next generation sequencing on positive specimens.

We estimated influenza VE using a test-negative design, comparing the odds of vaccination between influenza test-positive cases and test-negative controls identified during June 2021 through May 2023.(4) Influenza seasons in Thailand are defined as June through May of the following year.(2) We used a logistic regression model adjusted for age, sex, self-reported underlying medical condition, and calendar week.(5) VE was estimated as (1-adjusted odds ratio) * 100%. Secondary analyses were stratified by age group and by underlying medical conditions and were adjusted for sex and calendar week.

Overall, 11,312 patients had ILI or SARI during June 2021 through May 2023. After excluding 2,631 patients for having a positive SARS-CoV-2 result (n=2,600) or receiving influenza vaccination <14 days before symptom onset (n=31), there were 8,681 patients included.

After notably low influenza circulation during the 2021–2022 season (10 influenza test-positive cases), influenza cases increased at the beginning of the 2022–2023 season (Figure). Cases peaked at the end of August (predominantly A/Darwin/9/2021(H3N2)-like virus, clade 3C.2a1b.2a.2) with a second, smaller peak in late January 2023 (predominantly B/Austria/1359417/2021 [B/Victoria lineage]-like virus, subclade V1A.3a.2). Vaccination coverage was 5.6% of all included patients in 2021–2022 and 7.0% in 2022–2023.

**Figure.**
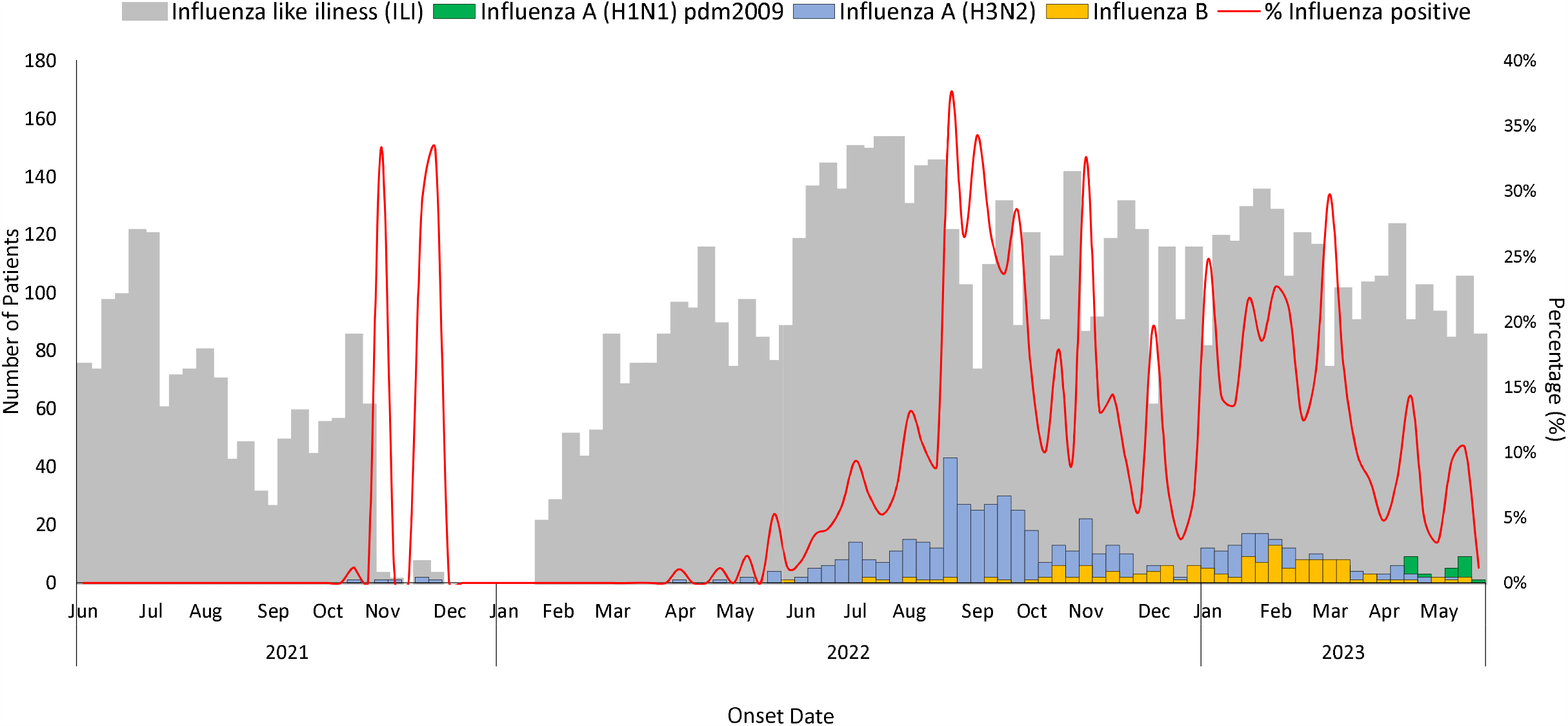
Influenza virus circulation by subtype and lineage from influenza sentinel surveillance during 2021–2022 and 2022–2023 seasons in Thailand.

VE was not calculated for 2021–2022 because there were too few influenza cases. In 2022–2023, adjusted VE against any medically attended illness (ILI or SARI) was 43% (95% confidence interval [CI] 15–61) (Table). Against influenza A, VE was 38% (95% CI 5–69) and against influenza B, VE was 61% (95% -6–86). Influenza VE against outpatient visits was 52% (95% CI 23–70), but we were unable to obtain a stable VE estimate against influenza hospitalization (VE 24%, 95% CI -55–63). Influenza vaccine was effective against any medically attended illness for adults ages 18–64 years (61%, 95% CI 24–80) and for individuals without underlying medical conditions (45%, 95% CI 21–66).

**Table.** Influenza adjusted vaccine effectiveness during June 2022–May 2023 influenza season in Thailand.

|  | Influenza test-positive cases<br>no. | Vaccinated<br>cases<br>no. (%) | Influenza test-negative<br>controls<br>no. | Vaccinated<br>controls<br>no. (%) | Adjusted vaccine<br>effectiveness*<br>(95% confidence<br>interval) |
| --- | --- | --- | --- | --- | --- |
| **Overall medically attended** |  |  |  |  |  |
| **Illness** |  |  |  |  |  |
| Influenza (any) | 768 | 29 (3.8) | 5,107 | 382 (7.5) | 43 (15, 61) |
| Influenza A | 622 | 25 (4.0) | 5,107 | 382 (7.5) | 38 (5, 69) |
| A (H3N2)† | 549 | 19 (3.5) | 5,107 | 382 (7.5) | 44 (9, 65) |
| A (H1N1)pdm2009‡ | 73 | 6 (8.2) | 5,107 | 382 (7.5) | 5 (-125, 60) |
| Influenza B§ | 146 | 4 (2.7) | 5,107 | 382 (7.5) | 61 (-6, 86) |
| **Secondary stratified analyses** |  |  |  |  |  |
| **Outpatient visit** |  |  |  |  |  |
| Influenza (any) | 550 | 20 (3.6) | 2,645 | 237 (9.0) | 52 (23, 70) |
| Influenza A | 456 | 17 (3.7) | 2,645 | 237 (9.0) | 49 (15, 70) |
| A (H3N2) | 413 | 13 (3.2) | 2,645 | 237 (9.0) | 55 (21, 75) |
| A (H1N1)pdm2009 | 43 | 4 (9.3) | 2,645 | 237 (9.0) | 10 (-162, 69) |
| Influenza B | 94 | 3 (3.2) | 2,645 | 237 (9.0) | 64 (-16, 89) |
| **Hospitalization** |  |  |  |  |  |
| Influenza (any) | 218 | 9 (4.1) | 2,462 | 145 (5.9) | 24 (-55, 63) |
| Influenza A | 166 | 8 (4.8) | 2,462 | 145 (5.9) | 15 (-82, 60) |
| A (H3N2) | 136 | 6 (4.4) | 2,462 | 145 (5.9) | 17 (-98, 66) |
| A (H1N1)pdm2009 | 30 | 2 (6.7) | 2,462 | 145 (5.9) | 3 (-332, 78) |
| Influenza B | 52 | 1 (1.9) | 2,462 | 145 (5.9) | 58 (-215, 94) |
| **6 months–17 years** |  |  |  |  |  |
| Influenza (any) | 586 | 14 (2.4) | 3,733 | 154 (4.1) | 34 (-17, 63) |
| Influenza A | 476 | 13 (2.7) | 3,733 | 154 (4.1) | 21 (-43, 57) |
| A (H3N2) | 422 | 11 (2.6) | 3,733 | 154 (4.1) | 19 (-56, 58) |
| A (H1N1)pdm2009 | 54 | 2 (3.7) | 3,733 | 154 (4.1) | 28 (-202, 83) |
| Influenza B | 110 | 1 (0.9) | 3,733 | 154 (4.1) | 78 (-62, 97) |
| **18–64 years** |  |  |  |  |  |
| Influenza (any) | 164 | 10 (6.1) | 1,032 | 148 (14.3) | 61 (24, 80) |
| Influenza A | 128 | 7 (5.5) | 1,032 | 148 (14.3) | 64 (21, 84) |
| A (H3N2) | 111 | 4 (3.6) | 1,032 | 148 (14.3) | 75 (32, 91) |
| A (H1N1)pdm2009 | 17 | 3 (17.7) | 1,032 | 148 (14.3) | 2 (-253, 73) |
| Influenza B | 36 | 3 (8.3) | 1,032 | 148 (14.3) | 51 (-64, 85) |
| **65 years and older** |  |  |  |  |  |
| Influenza (any) | 18 | 5 (27.8) | 342 | 80 (23.4) | -68 (-409, 44) |
| Influenza A | 18 | 5 (27.8) | 342 | 80 (23.4) | -68 (-409, 44) |
| A (H3N2) | 16 | 4 (25.0) | 342 | 80 (23.4) | -42 (-379, 58) |
| A (H1N1)pdm2009 | 2 | 1 (50.0) | 342 | 80 (23.4) | -409 (-8,916, 71) |
| Influenza B | 0 | 0 | 342 | 80 (23.4) | NA |
| **Underlying medical conditions** |  |  |  |  |  |
| Influenza (any) | 93 | 10 (10.8) | 1,015 | 168 (16.6) | 36 (-28, 68) |
| Influenza A | 72 | 8 (11.1) | 1,015 | 168 (16.6) | 32 (-46, 68) |
| A (H3N2) | 62 | 6 (9.7) | 1,015 | 168 (16.6) | 39 (-46, 75) |
| A (H1N1)pdm2009 | 10 | 2 (20.0) | 1,015 | 168 (16.6) | -5 (-412, 78) |
| Influenza B | 21 | 2 (9.5) | 1,015 | 168 (16.6) | 46 (-134, 88) |
| **No underlying medical conditions** |  |  |  |  |  |
| Influenza (any) | 675 | 19 (2.8) | 4,092 | 214 (5.2) | 45 (21, 66) |
| Influenza A | 550 | 17 (3.1) | 4,092 | 214 (5.2) | 39 (-1, 73) |
| A (H3N2) | 487 | 13 (2.7) | 4,092 | 214 (5.2) | 44 (1, 69) |
| A (H1N1)pdm2009 | 63 | 4 (6.4) | 4,092 | 214 (5.2) | 12 (-151, 69) |
| Influenza B | 125 | 2 (1.6) | 4,092 | 214 (5.2) | 69 (-25, 92) |

\*Vaccine effectiveness was estimated using the test-negative design as 100% $\times (1 - \text{adjusted odds ratio})$ in which the adjusted odds
ratio represents ratio of odds of being vaccinated among influenza-positive cases to odds of being vaccinated among influenza-negative controls, adjusted for age, sex, calendar week, and self-reported presence of one or more underlying medical condition.
<sup>†</sup>Predominantly A/Darwin/9/2021 (H3N2)-like virus, clade 3C.2a1b.2a.2. <sup>‡</sup>A/Sydney/5/2021 (H1N1)pdm09-like virus, clade
6B.1A.5a.2a. <sup>§</sup>Predominantly B/Austria/1359417/2021 (B/Victoria lineage)-like virus, subclade V1A.3a.2.

Influenza virus circulation resumed with a peak around September 2022 and a small peak in January 2023, which is typical for Thailand.(2, 6) Adjusted VE estimates against any influenza in this analysis of 2023 Southern Hemisphere seasonal influenza vaccine were similar to unpublished estimates from South Africa and Peru.(7, 8) Adjusted VE estimates against hospitalization were also similar to recent estimates from five South American countries.(9) Small differences in estimates could be related to differences in study sample size, health seeking behavior, population characteristics, or circulating influenza strains. VE against hospitalization was difficult to estimate because very few vaccinated cases required hospitalization. Nevertheless, this analysis demonstrates that influenza vaccine provided moderate protection against influenza virus infection in Thailand.

## Data Availability

All data produced in the present study are available upon reasonable request to the authors

## Acknowledgments

We thank all surveillance officers for their contributions to this project. We also thank Drs. Eduardo Azziz-Baumgartner and Michael Jhung for their expert review.

